# Integrating Macroeconomic and Public Health Impacts in Social Planning Policies for Pandemic Response

**DOI:** 10.1101/2025.01.21.25320900

**Authors:** Ofer Cornfeld, Kaicheng Niu, Oded Neeman, Michael Roswell, Gabi Steinbach, Stephen J. Beckett, Yorai Wardi, Joshua S. Weitz, Eran Yashiv

**Author notes:** Corresponding authors: YW –; JSW –; EY –.

## Abstract

Infectious disease outbreaks with pandemic potential present challenges for mitigation and control. Policymakers make decisions to reduce disease-associated morbidity and mortality while also minimizing socioeconomic costs of control. Despite ongoing efforts and widespread recognition of the challenge, there remains a paucity of decision tool frameworks that integrate epidemic and macroeconomic dynamics. Here, we propose and analyze an econo-epidemic model to identify robust planning policies to limit epidemic impacts while maintaining economic activity. The model couples epidemic dynamics, behavioral change, economic activity, and feasible policy plans informed by respiratory disease threats of pandemic concern. We compare alternative fixed, dynamic open-loop optimal control, and feedback control policies via a welfare loss framework. We find that open loop policies that adjust employment dynamically while maintaining a flat epidemic curve in advance of the uncertain arrival of population-scale vaccination outperform fixed employment reduction policies. However, open loop policies are highly sensitive to misestimation of parameters associated with intrinsic disease strength and feedback between economic activity and transmission, leading to potentially significant increases in welfare loss. In contrast, feedback control policies guided by open loop dynamical targets of the time-varying reproduction number perform near-optimally when parameters are well-estimated, while significantly outperforming open loop policies whenever disease features and population-scale behavioral response are misestimated – as they inevitably are. These findings present a template for integrating principled economic models with epidemic scenarios to identify vulnerabilities in policy responses and expand policy options in preparation for future pandemics.

## 1 Introduction

The COVID-19 pandemic caused more than 700 million documented cases and more than 7 million documented fatalities worldwide between 2020-2023 [1]. The actual number of infections and fatalities has been far higher, e.g., total infections are unknown but likely in the billions, while extrapolation from subnational level reports of excess mortality suggest that > 20 million individuals died of COVID-19 from 2020-2022 [2, 3]. This catastrophic level of impact was anticipated, in part, through assessment of early data on outbreak strength and severity in the Wuhan province in China [4]. In the absence of immediate and effective interventions, infectious individuals could generate ℛ_0_ ∼ 3 secondary infections [5] with infection fatality rates of ∼ 0.5–0.8% [6]. Together, mathematical models of epidemic dynamics combined estimates of human-to-human transmission and age-dependent hospitalization/fatality risk [7, 8] to project cumulative fatality rates on the order of 60 per 100,000 in the absence of large-scale interventions – equivalent to ∼ 2M fatalities in the United States and ∼ 300K in the United Kingdom [4].

In response to the potential catastrophic threat of COVID-19, governments rapidly imposed social distancing and/or lockdowns to reduce contacts between susceptible and infectious individuals (including those who may be unaware they are infected [9]) as a means to reduce rates of new infections [10–12]. As but one example, model-inferred estimates suggest that ∼ 3.1 million deaths were averted in 11 European countries between February-May 2020 due to national lockdowns [13]. Likewise, related analysis of social distancing policies in China, South Korea, Italy, Iran, France, and the United States estimated that policy directives and restrictions led to more than 61 million averted cases between February-April 2020 [14]. However, such counterfactuals come with significant caveats. First, baseline epidemic models do not typically integrate behavioral change (and/or alternative mitigation steps) that could lead to reduction in transmission and fatalities [15]. Second, even if early transmission is averted, subsequent relaxation of policies can lead to rapid resurgence of infections and fatalities, e.g., China’s late 2022 re-opening likely led to more than 1.4 million fatalities in a 3 month period between December 2022 and February 2023 [16]). Third, societal scale lockdowns impose health costs, decreasing the frequency of regular clinical care visits including screening for cancers [17], while increasing social isolation that impacts the mental health of children and adults [18].

Local, regional, and national lockdowns also come with substantive socioeconomic costs. In macroeconomic terms, lockdowns reduce economic activity due to production declines and decreases in productivity, losses of revenue, and business closures that ripple across different economic sectors. People and policymakers are still dealing with the aftermath of lockdowns. For example, lockdowns are hypothesized to have driven a burst of inflation [19] driven, in part, by supply-chain disruptions. Likewise, changes in the labor market induced by the pandemic, including increases in remote work, shifts in jobs, industries, and occupations employment pattens, and shifts in market demand have continued to impact economic productivity and gross domestic product worldwide [20–22]. In a March 8, 2024, public event at the London School of Economics [23], the Federal Reserve Bank of New York President John Williams expressed dissatisfaction with econo-epidemic models. Despite significant effort, the failure to integrate principled economic models with epidemic scenarios generates vulnerabilities in policy responses that prioritize one of health or economic outcomes at the expense of the other. As a result, there are unresolved questions on the link between epidemic dynamics, policy response, and economic impacts spanning increases in inflation, supply chain dilemmas, and changes in the labor market [19–22].

This paper addresses the lacuna between public health policies that aim to decrease the morbidity and mortality associated with disease outbreaks, and social planning policies that aim to stimulate and sustain economic activity. In doing so, we integrate both sets of goals in a common valuation framework and ask: what feasible policies minimize health impacts while maximizing economic activity? To address this question, we develop a social planning policy analysis framework that (i) includes realistic, feasible policy plans that account for lags in implementation and discrete policy periods; (ii) utilizes a common ‘value of reduced mortality risk’ (VRMR) framework for jointly evaluating the macroeconomic and public health effects of policy objectives – VRMR quantifies the equivalent substitution between averted deaths and money [24]; (iii) accounts for behavioral response and the lack of precise information on (re)emerging diseases. By integrating a common valuation framework and iteratively updating policies through commonly measured indicators of disease impact, we explore when and how policy planners can feasibly achieve *nearly* optimal population-scale epidemic and economic objectives in the face of persistent uncertainty regarding transmission and responses at individual scales.

## 2 Results and Discussion

### 2.1 Econo-Epidemic Modeling framework

We developed an integrated econo-epidemic modeling framework amenable to a social planning problem that can be used to identify ‘optimal’ policies given variation in disease transmission, behavioral response, and economic output (Figure 1). To do so, we utilize a Susceptible-Exposed-Infectious-Recovered/Removed (SEIR) epidemic modeling framework to represent disease spread at population scales (full equations in Supplementary Information (SI) Text A). The time varying incidence, *β*_*t*_*SI*, given the susceptible fraction *S* and infectious fraction *I* is modulated by the transmission rate

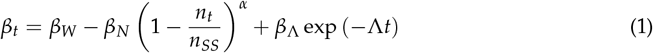

which is driven by a combination of factors: (i) baseline interactions when the economy is open (*β*_*W*_ + *β*_Λ_); (ii) rapid behavioral adaptation of the population over a short time scale (1/Λ) uncoupled to employment levels that reduces transmission by *β*_Λ_; (iii) endogenous behavioral response arising from individual-level decisions not necessarily mandated by policy (e.g., working from home, masking, and improved ventilation in the case of respiratory diseases); (iv) policy-induced dynamic reduction in transmission. The realized employment level *n*_*t*_ relative to the steady-state economy *n*_*SS*_ leads to a reduction in transmission parameterized by *β*_*N*_ and an exponent *α*. The resulting time-dependent effective reproduction number is therefore 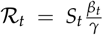, where *γ* is the removal rate of infectious individuals.

**Figure 1:**
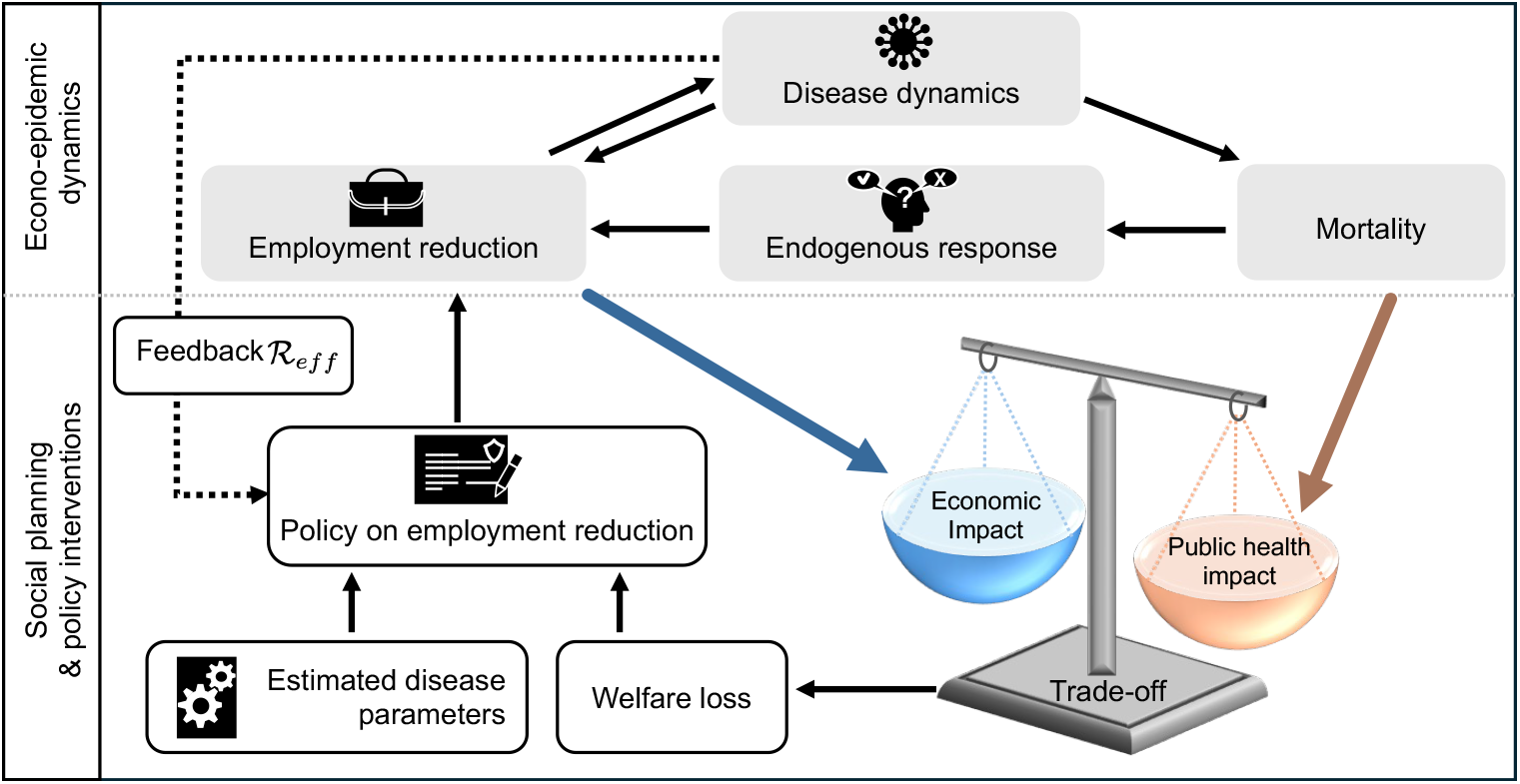
Schematic of the econo-epidemic modeling framework with epidemic dynamics, macroeconomic model of welfare loss, endogenous response, and feedback with a social planning problem. The top section provides an overview of the underlying econo-epidemic model. The bottom section provides an overview of the social planning process that integrates economic and public health impact as a means to develop optimal policies (both open- and closed-loop) to minimize welfare loss through modification of employment reduction policies. Full specification of the epidemic model dynamics, economic model structure, control theoretic approach, policy planner optimization, and disease parameters are found in Supplementary Text A.

The disease model is coupled to a macroeconomic model in which the gross domestic product (GDP) is driven by a linear production function tied to employment, assuming constant wages and that output is fully consumed (see SI Text A). Employment is influenced by (i) individual-level economic activity guided by utility maximization linked to the severity of the disease outbreak, 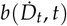, which we refer to as the endogenous behavioral response, and (ii) a social planner that imposes a level of preferred employment reduction, *L*_*t*_. We assume that the realized employment reduction is the maximum of these two effects, i.e.,

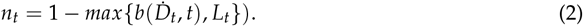

In this combined econo-epidemic modeling framework, the objective of the central planner is to minimize welfare loss *WL*(*L*_*t*_) caused by the disease, balancing the death toll with the economic costs (i.e., cumulative work hours, see SI Text A). Welfare loss is a function of the policy *L*_*t*_ equivalent to the fractional employment reduction – which typically exceeds the endogenous response. Welfare loss is measured by economic utility/welfare units and is nonlinearly related to wages, level of economic utility, the death toll, and the disutility from working. Throughout, we consider the social planning problem over a time horizon *T* in which we expect the large-scale dissemination of effective vaccines (Supplementary Table S5).

### 2.2 Evaluation of fixed employment reduction policies to minimize welfare loss during a pandemic

In the absence of social planning interventions, an initially small fraction of infected individuals will catalyze an outbreak leading to transient reduction in employment due to utility maximization via the endogenous behavioral response. This scenario (Supplementary Figure S1) leads to large-scale outbreaks and significant loss of life. It also provides the baseline for evaluating alternative, fixed economic reduction policies intended to reduce welfare loss. To generate the baseline dynamics associated with basic reproduction number ℛ_0_, we consider variation in fixed employment reduction policies across a continuum ranging from fully open to a maximally restricted economy (econo-epidemic model parameterization in Table S5). Employment reduction reduces cumulative fatalities while increasing economic loss. We identify an optimal, intermediate lockdown level corresponding to a fixed policy that minimizes welfare loss compared to viable alternatives (see minima in welfare loss in the top panels of Figure 2). Variation in either the intensity of baseline transmission (due to differences in disease features leading to changes in ℛ_0_) or the efficacy of employment reduction on transmission lead to different optimal, fixed employment reduction policies (Figure 2 bottom). Typically, increases in disease intensity and/or decreases in the efficacy of employment reduction on transmission increase the welfare loss associated with fixed, optimal policy responses. Hence, insofar as disease intensity and the link between employment reduction and transmission are known with certainty, there exists an optimal *fixed* response that can be planned in advance.

**Figure 2:**
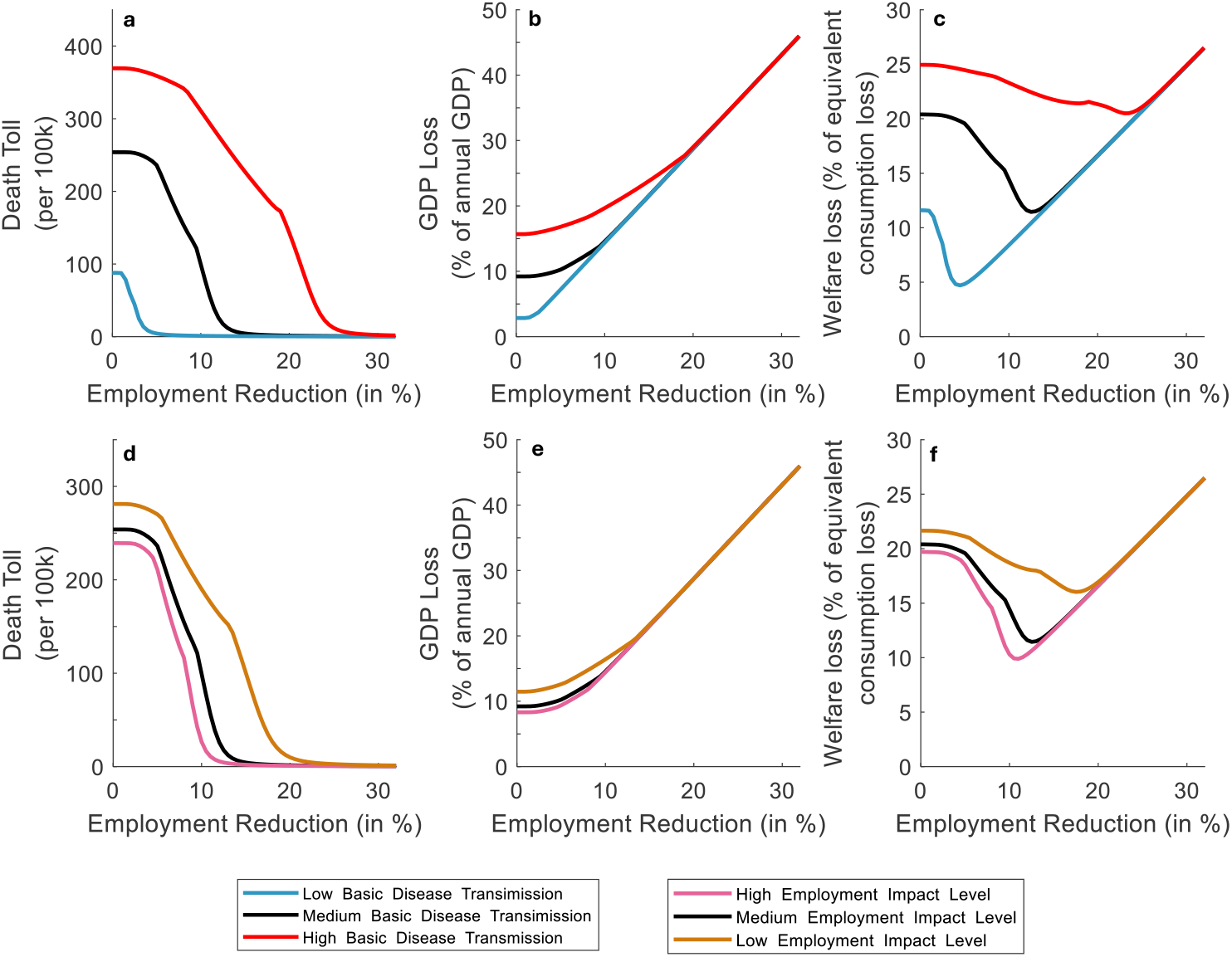
Optimal fixed employment reduction policies reduce welfare loss, balancing impacts of cumulative fatalities and reduction in GDP. Panels (A)-(C) show the outcome of fixed employment reduction (on the x-axis) in terms of fatalities (per 100,000), GDP loss (%), and % welfare loss), respectively. The three curves in each panel denote outcomes given baseline (black, ℛ_0_ = 2.86, /5yv = 0.376), elevated (red,ℛ_0_ = 3.156, fin = 0.45), or reduced (blue,ℛ_0_ = 2556, fiw = 0.3) disease transmission conditions, consistent with variation in early estimates of COVID-19 strength [5] The minimal WL* associated with the optimal, fixed employment reduction policy, n^*^ corresponds to the point at which welfare loss is at its minimum. Likewise, Panels (D)-(F) show modulation of the impact of employment reduction on transmission using the disease parameters in the black curve conditions in Panels (A)-(C), given more impactful (pink, *β*_N_ = 0.6), and less impactful (orange, *β*_N_, = 0-4) conditions. The baseline employment impact is when *β*_N_ = 0.53.

### 2.3 Open loop control policies outperform fixed lockdown policies in minimizing welfare loss during pandemics

We sought to identify and characterize optimal open loop, *time-dependent* policies given continuous variation in employment reduction levels *n*_*t*_, rather than fixed employment reduction policies as explored in the previous section. To do so, we pose and solve an optimal control problem using a robust steepest-descent algorithm based on the maximum principle (detailed in the Supplementary Information). We utilize the employment reduction level, *n*_*t*_, as the control variable accessible by the policy maker which influences transmission and the effective reproduction number. Figure 3 panels (a-c) and (d-f) compare consequences for welfare loss and optimal control solutions *n*_*oc*_(*t*) for low, medium, and high basic transmission cases, spanning ℛ_0_ ≈ 2.6, 2.9, and 3.2 respectively. In each case, the optimal control algorithm identifies time-dependent changes in employment reduction (see Figure S2 for disease dynamics, ℛ_*eff*_, and welfare loss). Initially, the economy is restricted with significant economic cost. Given low prevalence (and low mortality), the rapid learning period reduces transmission (e.g., via masks, social distancing, and crowd avoidance), leading to a reduction of ℛ_*eff*_ close to, but slightly above 1. Then, exponential increases in disease burden drives a second phase of reduced employment that exceeds employment reduction expected through the endogenous response alone. Hence, the open loop optimal control policy reduces ℛ_*eff*_ slightly below 1. Finally, the expected arrival of an effective vaccine disseminated at high coverage allows the optimal planner a means to reduce restrictions. These three phases appear most evidently in the high disease scenario, but are present in each of the low, medium, and high transmission scenarios in the optimal continuous policy. The equivalent total welfare loss for the optimal *time-dependent* policy is shown as a function of ℛ_0_ in panels (a)-(c). We also confirm that these time-dependent policies could be implemented feasibly, i.e., by restricting the interval length during which a policy could be changed. In practice, we offer the planner limited flexibility, showing that 3 policy regimes are sufficient for an 18-month intervention period. The optimal piecewise constant curves (i.e., ‘optimal stepwise policies’) are paired with each optimal continuous policy in panels (d)-(f), closely mimicking the optimal continuous policy both in shape and in performance. Notably, the optimal time-dependent policies (whether continuous or stepwise) each identify nearly the same level of employment reduction. However, the time at which the optimal control algorithm identifies the appropriate moment to shift between policies (initial, restricted, relaxed) varies with the underlying disease strength (see Figure S2). This variation also suggests that misestimation of disease strength during the planning policy could lead to mismatched responses.

**Figure 3:**
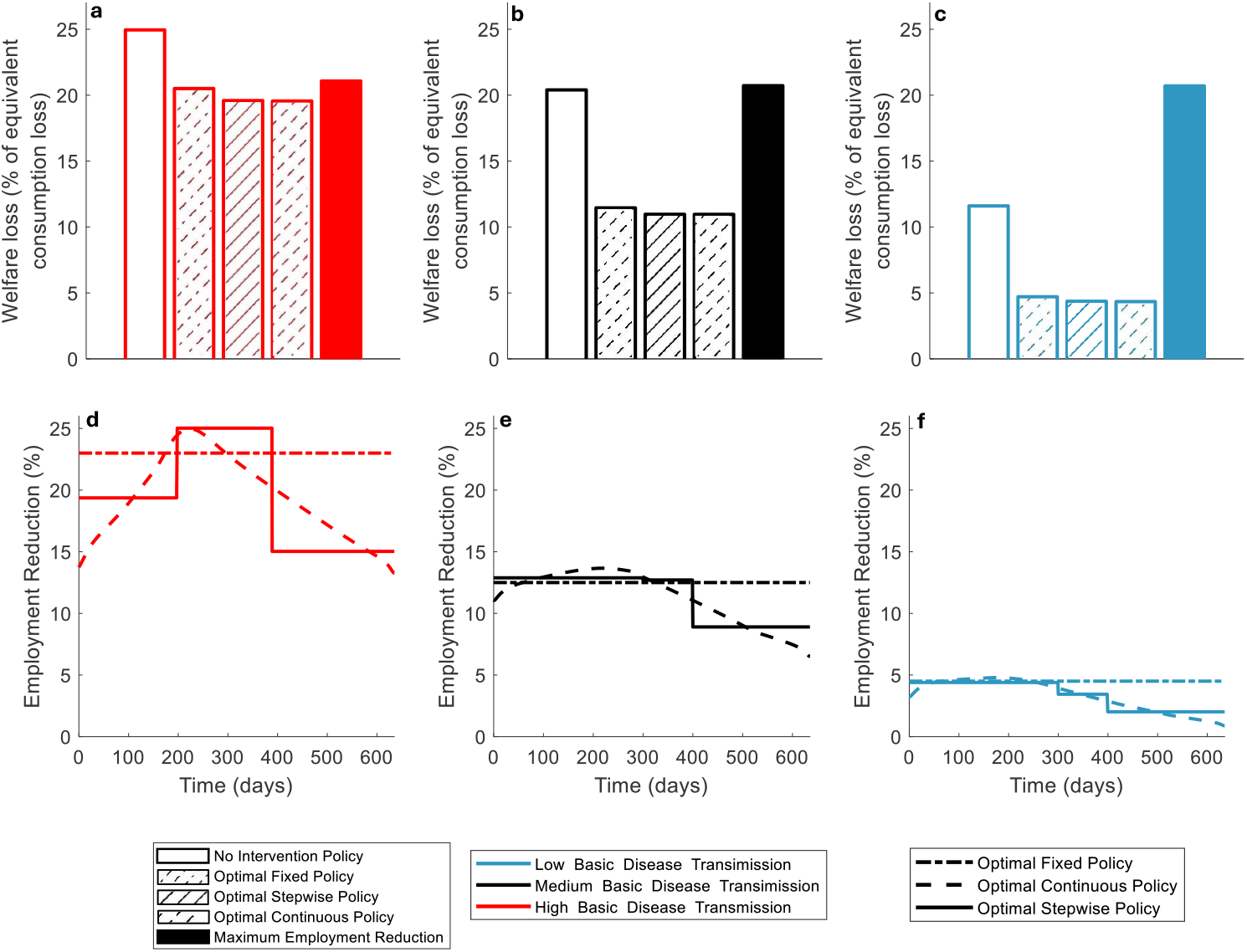
Performance of open loop, optimal control policies across disease transmission conditions. Three optimal poli-cies are contrasted with no intervention and maximal intervention options. The optimal policies include an optimal fixed,continuous, and stepwise policy. Panels (a)-(c) show the cumulative welfare loss for all five cases, in each case the optimalpolicies outperform either no intervention or full restrictions. Panels (d)-(f) show the employment reduction over time forthe three optimal policies. Each of the three plots in the 2 panels denotes the policies and outcomes for 3 different diseasetransmission conditions: Low (ℛ_0_ = 2.56), medium (ℛ_0_ = 286), and high (ℛ_0_ = 3.16). Across conditions, the optimalstepwise policy closely resembles the continuous optimal policy, and consistently outperform fixed policies.

### 2.4 Fragility of optimal control policies given uncertainty

Optimal control problems can be sensitive to misspecification of parameters, especially when applied to nonlinear dynamic systems with the potential for (transient) exponential growth [25]. Hence, we set out to evaluate the sensitivity of performance, as measured in terms of welfare loss, given solutions of the optimal control algorithm for parameters *θ*_*ref*_ when the disease outbreak is characterized by 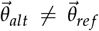. As above, the optimal control problem is solved using a model-based, open loop, offline computation yielding time-dependent policies for transmission that can be mapped to equivalent employment reduction policies *n*_*t*_. Figure 4 highlights the sensitivity of the optimal *time-dependent* policy to misspecification of parameters. The purple curves in panels (b)-(d)show the difference between the death toll, GDP loss, and welfare loss relative to the optimal time-dependent policy given a reference basic reproduction number (x-axis, vertical dashed line). When the pathogen is less transmissable, then the optimal policy will be overly cautious, leading to modest decreases in the death toll, substantial increases in GDP loss, and substantial increases in welfare loss, jut as fixed policies are prone to misspecification errors (as in Figure 2). Likewise, when the pathogen is more transmissable, then the optimal policy will be insufficiently cautious, leading to substantial increases in the death toll, modest improvements in GDP loss, and substantial increases in welfare loss (Figure 2).

**Figure 4:**
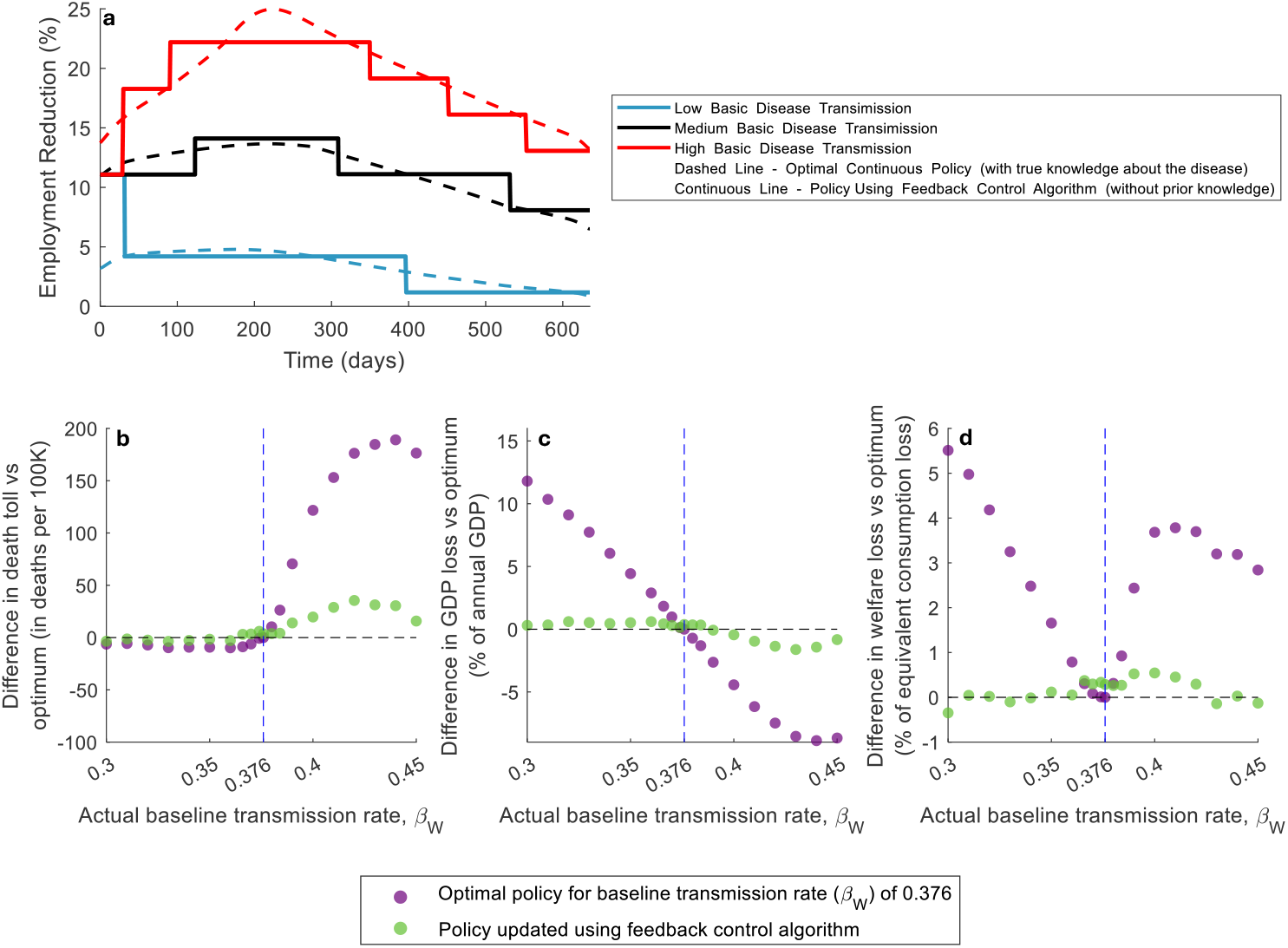
Comparison of epidemic outcomes given social planning policy guided by feedback control vs. open loop control, (a) Employment reduction policies in the case of low, medium, and high disease transmission respectively. The policies contrast the optimal continuous policies given accurate knowledge of the disease state (dashed line) with a feedback control policy that does not use direct information about the disease state (solid line). Impact of mis-specification of disease parameters given variation in the basic transmission level, *β*_W_ given differences in (b) death toll, (c) GDP loss, and (d) relative change in WL. The optimal policy is highly sensitive to misspecification of disease transmission rates, whereas the feedback control policy approach is not. The feedback policy maintains low levels of death, GDP loss, and overall welfare loss across different estimates of disease strength during planning.

### 2.5 Robust feedback control in econo-epidemic models

Identifying optimal,time-dependent planning policies via open loop algorithms leads to improvements in welfare loss compared to fixed policies (see Figure 3) provided they rely on accurate disease parameter estimates (see Figure 4). However, a comparison of optimal time-dependent policies did yield a dynamical insight – despite differences in employment reduction associated with variations in underlying disease strength, the target levels of ℛ_*eff*_ were relatively robust. For example, when varying ℛ_0_ from 2.556 to 3.156, while we found approximately 400% relative differences in employment reduction during the restricted phase, the ℛ_*eff*_ relative difference was around 6% (See Figure S2). In this restricted phase, we observe an emergent feature of disease transmission dynamics – the disease is controlled at levels where ℛ_*eff*_ < 1, but only slightly so. Maintaining the effective reproduction number below 1 constrains exponential increases in incidence without paying the economic cost of more restrictive measures. Hence, we implemented a feedback control planning algorithm that tracks ℛ_*eff*_ (note that real-time estimates of the effective reproduction number are increasingly accessible [26]). We implement the feedback control using the proportional-integral-derivative (PID) control technique [27] (the algorithm is detailed in Supplementary Text B.2). Figure 4a specifies the resulting feedback control policy when optimized for the correct and mismatched disease parameters (both stronger and weaker than the reference parameters). Note that despite the mis-specification, the feedback control policy identifies similar (albeit slightly lagged) shifts in the timing between initial, restricted, and relaxed phases. Moreover, the welfare loss under the feedback control policy is robust to mis-specification of parameters (also see robustness with respect to the link between employment and transmission in Figure S4). This robustness of policy response contrasts with the extreme sensitivity of the optimal time-dependent employment reduction policy identified through an open loop optimal control algorithm (contrast green, feedback control with purple open loop, optimal control in Figure 4d).

## 3 Discussion

We developed and analyzed a social planning problem centered on an econo-epidemic model that couples transmission dynamics between individuals with changes in employment. Our objective was to identify a suite of feasible and robust social planning policies that could minimize welfare loss as measured in terms of the value of reduced mortality risk, i.e., accounting for fatalities averted during the pandemic as well as GDP decreases arising from employment reduction. In doing so, we considered a fully coupled model such that changes in disease severity would decrease employment through endogenous feedback which, in turn, would lead to decreases in transmission. The social planner then has the opportunity to go beyond endogenous response and restrict economic activity. As we show, although it is possible to devise an optimal, dynamic policy with reduced employment that outperforms any fixed policy (e.g., lockdowns or otherwise), such optimal dynamic policies can be extremely sensitive to misestimation of disease transmission parameters and/or the impact of economic activity on disease transmission. Indeed, implementing the incorrect ‘optimal’ dynamic policy can lead to mismatched timing of interventions and significant increases in welfare loss. Instead, we show that such optimal dynamic policies can be used as a guide for a feedback control policy, leveraging robustness properties and implementation principles of proportional-integral-derivative (PID) controllers. As a result, a social planner can implement a feedback control policy that is feasible (i.e., is implemented via a combination of fixed policy blocks), nearly-optimal (i.e., performs nearly as well as the optimal dynamic policy with perfect information), and robust to mis-specification (i.e., continues to perform nearly as well as the optimal dynamic policy even when parameter estimations are misaligned with reality). If prepared in advance, such social planning policies could counter false dichotomies surrounding prioritization of public health or the economy.

The COVID-19 pandemic is unlikely to be the last. Increasing mobility that enables long-distance transmission, changes in climate that facilitate expansion of pathogen geographic ranges, and increasing stress placed at human-zoonotic interfaces can each contribute to increasing the pandemic potential of endemic and emerging pathogens. Specific threats include COVID-19, H5N1 (and other avian influenza variants), as well as vector-borne viruses with pandemic potential (Zika, Nipah, and others) [28–32]. These diseases pose an increasing and critical threat to global health and economic security. The June 2021 report of a high-level G20 panel posits that “We are in an age of pandemics. There is every likelihood that the next pandemic will come within a decade — arising from a novel influenza strain, another coronavirus, or one of several other dangerous pathogens. Its impact on human health and the global economy could be even more profound than that of COVID-19.” Hence, response to pandemic threats requires planning scenarios that address the joint problem of mitigating transmission risk while minimizing socioeconomic impacts. For example, a study preceding the COVID-19 pandemic estimated that pandemic impacts might approach 500 billion dollars per year (0.6% of global income) [33]. In fact, GDP decreased by ≈ 3% in 2020, or approximately 2.5 trillion dollars [34], consistent with interquartile range estimates of 2.6%-4.2% total GDP loss per year due to global warming by 2050 under a 1.5^°^C increase scenario [35]. There is a clear need to leverage lessons learned from the COVID-19 response and improve public health infrastructure. However, social fatigue, the spread of misinformation, and politicization of public health response each presents challenges to coordinated responses if a novel threat were to arise.

Here, the social planning response is guided by an idealized model of disease spread coupled to an economic model. Both the economic and epidemic model come with caveats. The economic model is simplified. It can be extended by modeling the heterogeneity of individuals [36] and of firms or sectors [37], explicit modeling of costs to policy implementation, and the formulation of learning mechanisms [38]. Likewise the epidemic model includes a relatively simplified representation of outbreak dynamics. The model neglects differences in asymptomatic, presymptomatic, and symptomatic transmission, does not account for age-structure or heterogeneous mixing, stochasticity, evolution of strains, nor spatially explicit dynamics arising from a combination of long-distance travel and local mobility patterns. Nonetheless, the framework presented here could be adapted to variations of both the economic and/or epidemic components of the model. In doing so, it will be essential to consider to what extent social planning is feasible, improves upon expected endogenous responses to epidemics, and does not unintentionally induce increases in welfare loss.

Consistent with prior work focusing on control strategies to manage COVID-19 epidemic dynamics (in the absence of socioeconomic feedback [25, 39] we find that optimal dynamic control policies are highly sensitive to misspecification of dynamics, lead to mistimed interventions, and increases in welfare loss. Although feedback control policies are robust to the assumptions and feedback in the present econo-epidemic framework, it will be essential to evaluate robustness to structural and parameter uncertainty moving forward. Implementing policies that reduce welfare loss also depends on the extent to which individuals take steps to reduce interactions rationally in response to perceived risk of infection. Increasing polarization [40] could limit endogenous responses, thereby increasing the need for intervention policies, while at the same time undermining the effectiveness of policies. We recommend that efforts to communicate optimal feedback control policies prioritize communication of the benefits and rationale behind policies – both in terms of public health and socioeconomic benefits. Doing so will not just require development of more sophisticated models, but an increasing willingness to collaborate across social sciences, economics, and public health.

## Acknowledgments

We gratefully acknowledge funding from the Simons Foundation grant 930382 (to JSW) and support from the Chaires Blaise Pascal program of the Île-de-France region (to JSW). We thank M. Hochberg for feedback at an early stage of development of this manuscript, and participants at the Brin Mathematics Research Center workshop on Disease Dynamics and Human behavior that took place in November 2024 at the University of Maryland, College Park for useful conversations and feedback and the support of the Research Development Office at the University of Maryland for support in graphics development.

## Data availability statement

All code and simulation data is available at https://github.com/odedneeman/Optimal-Pandemic-Control and will be archived via zenodo.org.

## Supplementary Information for

### A Econo-Epidemic Model Framework

#### A.1 The Planner Objective Function

A central social planner aims to minimize welfare loss *WL* caused by the disease, balancing the death toll with economic cost. This is done by using a policy tool – employment reduction (*L*_*t*_) – which restricts economic activity in order to reduce the spread of the disease. By economic activity, we mean employment (hours of work), though this can be generalized.

The objective function of the controlled variable *L*_*t*_, to be minimized, is given by:

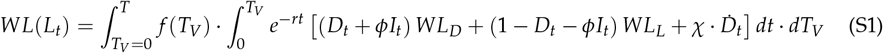

where

i. *f* (*T*_*V*_)− PDF of vaccine arrival time *T*_*V*_.
ii. 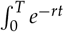− discounting of future values with interest rate *r*. Time *T* is the planner horizon.
iii. The fraction of the population (itself normalized to 1) who do not work (including the infected who are isolating) is given by *D*_*t*_ + *ϕI*_*t*_, where *WL*_*D*_ is the loss of welfare associated with a person not producing.
iv. The fraction of the population working is given by (1 − *D*_*t*_ − *ϕI*_*t*_) and *WL*_*L*_ is the loss of welfare associated with a production level below an endogenously preferred level (more on this below).

The contribution to welfare loss from the death toll is given by 𝒳 · 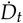 with 𝒳 > 0 is the value of reduced mortality risk and 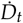 is the flow of deaths.

Welfare loss is measured by economic utility/welfare units, where we follow the modeling precedents of previous research [37]. For welfare itself we use 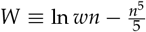 where *n* is the level of economic activity (fraction of daily hours worked), *w* is the daily wage, and 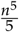 is the disutility from working, further explained below.

Specifically:

1. 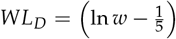 is the welfare from working full time; this is lost for people not working,*D*_*t*_ + *ϕI*_*t*_.
2. 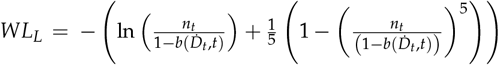 is the loss of welfare when employment *n*_*t*_ is restricted by policy to be below the level endogenously chosen by individuals, discussed below.
3. The level of economic activity is restricted by 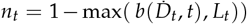, where *L*_*t*_ is the policy tool and 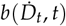, captures the endogenous response of individuals, which expresses both fatalities awareness (being a function, *b*, of the flow of deaths, 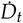) and issues related to time, such as fatigue (dependence on *t*).
4. Both *L*_*t*_ and 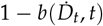, are bounded from above by 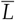 capturing the maximum attainable restriction on economic activity.

Planner’s horizon *T* is set so that the probability of vaccine arrival before that day exceeds 99%.

The idea of a social planner has been used extensively in studying social welfare. The latter concept has been discussed since the early 20th century (see for example [41–43]). A social planner is a hypothetical decision-maker who attempts to maximize some notion of social welfare. The planner is a fictional entity who chooses allocations for every agent in the economy that maximize a social welfare function subject to certain constraints. The welfare loss function used here serves this purpose and has been widely used in studying COVID 19, as for example in [44].

#### A.2 Epidemic Dynamics

We use the SEIR model framework while explicitly tracking deaths. The following equations describe the nonlinear dynamics of this model, in which each variable represents a fraction of the total population.

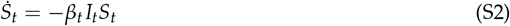

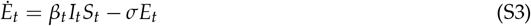

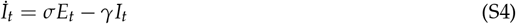

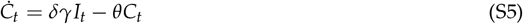

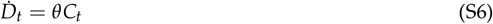

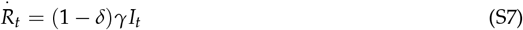

where *R* is the fraction of the infected that recover and *C* is the fraction who become severely sick and eventually die. Throughout, *β* denotes a transmission rate, *σ* is the incubation rate, *γ* is the removal rate of which 1 − *δ* recover and *δ* transition to a severely sick state leading to new fatalities given a death rate *θ* given severe illness. In this model, the basic reproduction number ℛ _0_ is given by:

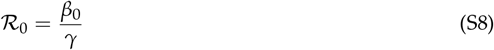

and the time-varying effective reproduction number is given by:

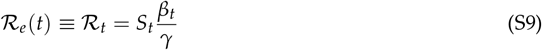

We model the transmission rate *β*_*t*_ as a function of three factors:

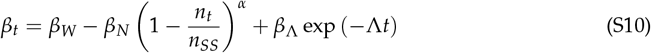

where *n*_*t*_ is the employment reduction relative to the maximum *n*_*SS*_. The formulation is motivated as follows:

a. *β*_*W*_ is the transmission rate when the economy is open, i.e., production and employment are not restricted.
b. *β*_*N*_ parametrizes the scale of the decline in transmission as activity falls (decline in 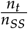, employment relative to its steady state), using a power function with parameter *α*.
c. *β*_Λ_ exp (− Λ*t*) expresses the decline in transmission due to rapid learning over time by individuals after the outbreak begins over a characteristic time scale 1/Λ.

Thus, at time *t* = 0, when 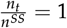 we get:

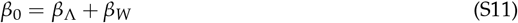

which is the transmission at the initial stage and corresponds to 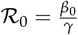.

After a period of time, which depends on the rate of decline Λ, individuals change their behavior, and when exp (− Λ*t*) ≪ *β*_*W*_ we get that the transmission rate (and hence ℛ_*t*_) rises with employment:

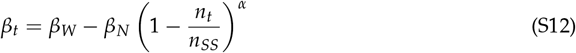

#### A.3 Macroeconomic model

The economy is modeled via a linear production function, with constant wages, and output is fully consumed, where all state variables are monitored per-day. In the model, *y*_*t*_ is GDP and *c*_*t*_ is consumption. The government imposes a lockdown policy *L*_*t*_. Steady state (SS) employment (fraction of daily hours worked) is 1, i.e., *n*_*SS*_ = 1. Total population is also normalized to 1. The following are the key relations:

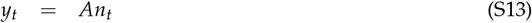

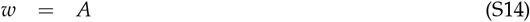

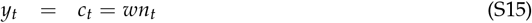

Next, we provide details on the factors that set employment *n*_*t*_.

**Individual Utility Maximization** Individual utility is given by:

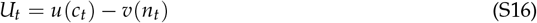

There is positive utility from consumption and disutility from labor. Prevalent functional forms are:

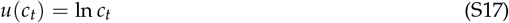

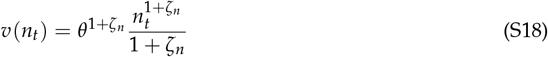

We use an empirically based value of Frisch elasticity ζ_*n*_ = 4. In the optimal solution 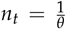 as 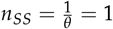, we get *θ* = 1.

At steady state where *n*_*SS*_ = 1 we get:

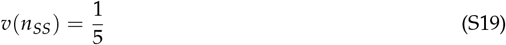

such that

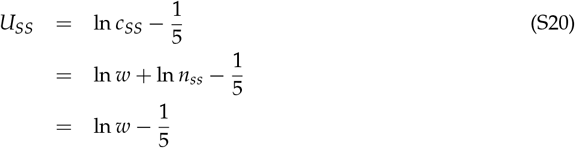

Out of steady state we have:

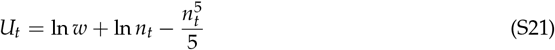

In the outbreak, there is an endogenous response of individuals to incident fatalities,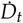, so the utility function is modified as follows:

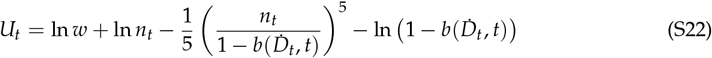

where *b* is the endogenous response.

**Consequences for Employment** The macroeconomic model assumes that employment is determined in two ways:

a. the planner imposes lockdowns *L*_*t*_. This is set by equation (S1), i.e., by minimizing social harm subject to all of the constraints.
b. the individual sets desired employment by utility maximization as discussed in sub-section A.3.

We posit that the stricter reduction in employment – by lockdown or by the individual response dominates. Employment thus behaves as follows:

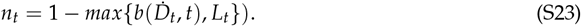

### A.4 Computing Welfare Loss In Equivalent Consumption Terms

We wish to convert welfare loss to equivalent consumption loss. Consider steady state and period *t* welfare. Using Eqs. S20 and S21 we define welfare loss *WL*_0_ (*n*_*t*_) to be:

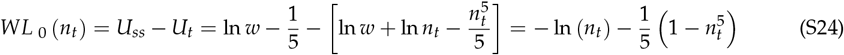

So the inverse function *WL*_0_ ^−1^ (*WL*_*t*_) converts welfare loss terms into an equivalent loss in employment and consumption terms.

For example *WL* _0−1_ (0) = 1, so *n*_*t*_ = 1. There is no reduction in welfare; this corresponds to full employment.

As *WL*_*t*_ rises *WL*_0_ ^−1^ (*WL*_*t*_) falls, so we define the corresponding employment and consumption reduction, *CL*, as:

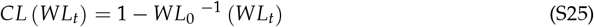

This function maps *WL*_*t*_ to the corresponding equivalent reduction in consumption (in %). In our model *WL*_*t*_ is the welfare loss, which is the integrand of the objective function :

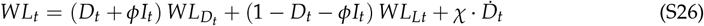

In order to compute the equivalent consumption terms loss we compute

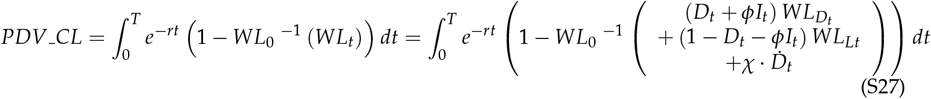

### B Optimal Policy Identification

#### B.1 Optimal control algorithm

In line with formalism from theories of optimal control [45], the state equation has the form

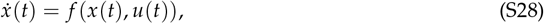

where *t* ∈ [0, *t* _*f*_] is the time-variable for a given *t* _*f*_ ∈ (0, ∞), *x*(*t*) ∈ ℝ ^*n*^ is the state variable and *u*(*t*) ∈ ℝ^*k*^ is the input-control variable. We assume that an initial condition *x*(0) := *x*_0_ ∈ ℝ^*n*^ is given and fixed. The optimal control problem is to compute a control { *u*(*t*) : *t* ∈ [0, *t* _*f*_] } which minimizes the following cost functional,

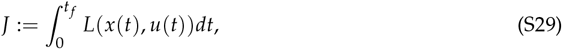

for a suitable cost function *L* : ℝ^*n*^*×* ℝ^*k*^ → ℝ, subject to pointwise constraints of the form *u*(*t*) ∈ 𝒰(*t*), where 𝒰(*t*) ⊂ ℝ^*k*^ is a time-dependent compact, convex set. Additional constraints on the input-control signal may be imposed such as piecewise continuity in the time-variable *t*. If the input control *u*(·) satisfies all of those requirements, it is said to be *admissible*. The costate variable *p*(*t*) ∈ ℝ^*n*^ is defined by the following equation,

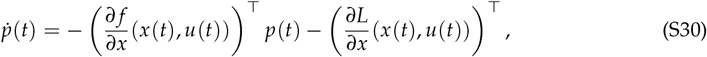

with the boundary condition *p*(*t* _*f*_) = 0. We remark that the boundary condition *p*(*t* _*f*_) is specified at the final time *t* _*f*_, and not at the initial time *t*_0_ := 0 as for the state equation (S28). Therefore, numerical computations of the costate have to be performed backwards in time after the state *x*(*t*) has been computed for all *t* ∈ [0, *t* _*f*_]. Throughout the forthcoming discussion, the time variable *t* ∈ [0, *t* _*f*_] is continuous. The term “computation of a time-dependent variable for all *t* ∈ [0, *t* _*f*_]” implicitly assumed computation over a given approximation grid.

A key element in the characterization of an optimal control is the Hamiltonian function *H* : ℝ^*n*^ *×* ℝ^*k*^ *×* ℝ^*n*^ → ℝ, defined as

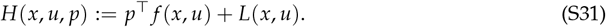

Given an admissible control *u*(·), let *x*(·) and *p*(·) be the state trajectory and costsate trajectory, respectively, associated with *u*(·). Given another admissible control, *v*(·), define the function 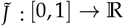 in the variable *λ* as

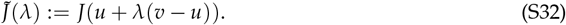

Then under suitable assumptions [46], the one-sided derivative 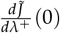 satisfies the following inequality,

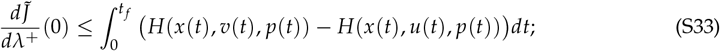

we note that here *x*(*t*) and *p*(*t*) are the state variable and costate variable, respectively, corresponding to the input control *u*(·). This result implies the maximum principle whereby if *u*(·) is an optimal control, then at almost every *t* ∈ [0, *t* _*f*_], *u*(*t*) is a minimizer of *H*(*x*(*t*), *v, p*(*t*)) over *v* ∈ 𝒰 (*t*). The reason for this is that if the maximum principle is not satisfied at *u*(·), then a descent direction for *J*(·) can be obtained from *u*(·) by *v*(·) satisfying the inequality

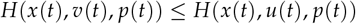

for all *t* ∈ [0, *t* _*f*_], and

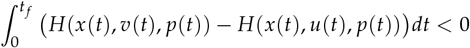

contradicting the supposition that *u*(·) is an optimal control. In the special case where *v*(*t*) is a pointwise minimizer of *H*(*x*(*t*), *v, p*(*t*)) for all *t* ∈ [0, *t* _*f*_], *v*(·) is a steepest descent direction.

The maximum principle leads us to the following iterative algorithm for the optimal control problem.

##### Hamiltonian-based algorithm

Fix a constant *λ* > 0 (to be used as a stepsize) and an initial control, *u*_0_(·).

Given a control *u*_*k*_(·), *k* = 0, 1, 2, …, compute the next control, *u*_*k*+1_(·), as follows.

*Step 1:* Compute (numerically, via an approximation) the state trajectory *x*_*k*_(·) defined by (S28), and the costate trajectory *p*_*k*_(·) defined by (S30).

*Step 2:* For each *t* ∈ [0, *t* _*f*_], compute a point *v*_*k*_(*t*) ∈ R^*k*^ satisfying

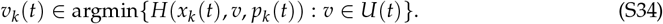

*Step 3:* Set *u*_*k*+1_(·) = *u*_*k*_(·) + *λ*(*v*_*k*_(·) − *u*_*k*_(·)).

For the step size we chose *λ* = 0.01. In making this choice we err on the side of simplicity of coding the algorithm while slowing down its convergence rate as compared to larger, variable step sizes without guarantees of convergence but a faster approach towards regions of optimal control. This choice was based on the realization that in a practical setting of pandemic management the optimal control program would be executed, off line, once per several days or weeks.

#### B.2 Feedback control algorithm

Due to the fragility of open-loop optimal control, it is a common practice in applications to first compute an open-loop optimal control solution, use it to compute a corresponding output, then use that output as the reference signal to be tracked in real time by the feedback control. Observe that the formulation of the optimal control problem defined by Eqs. S28 and S29 makes no reference to a system’s output, and it is up the the feedback-controller’s designers to choose an output according to practical considerations including the effectiveness and efficiency of the control law.

A schematic for the closed-loop system is depicted in Figure S3. The reference signal *r*(*t*) is the object of tracking by the output of the econo-epidemic system, *y*_*t*_. The input to the econo-epidemic system, *u*(*t*), is the output of the secondary controller, which has two inputs: the error signal *e*(*t*) = *r*(*t*) − *y*(*t*), and the output error *u*(*t*) − *u*_*c*_(*t*), the latter the output of the PID controller. The objective of the controller is to ensure that lim sup_*t*→ ∞_ ||*e*(*t*)|| be small to within specifications.

The optimal control problem considered in this paper has the particular constraint that the input signal *u*(*t*), representing a policy, be piecewise constant with a limited number of value-switchings and lower bounds on the dwell-times (i.e., lengths) of constant-value periods. This constraint classifies the problem in the category of hybrid switched-mode optimal control [47], whose solution, typically by computational means, may be complicated and time consuming due to the presence of large numbers of local minima. An alternative approach is to migrate the task of guaranteeing the constraint from the optimal-control’s algorithm to the design of the feedback control. What makes this approach reasonable is the fact that for the considered optimal control problem, a suitable choice of the system’s output, *y*(*t*) = ℛ_*t*_, computed from an optimal control solution, has a near-constant value throughout a large part of the time horizon for the problem. Furthermore, the value of that constant is robust with respect to tested model-parameter uncertainties.

The feedback control law that we chose is founded on a version of the Proportional-Integral-Derivative (PID) controller [27]. In continuous-time systems, the commonly-used PID control has the following form,

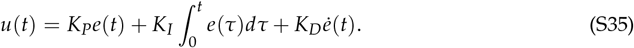

The designations “proportional”, “integral” and “derivative” refer to the three respective relationships between *e*(*t*) and *u*(*t*) defined by Eq. S35. Thus, the first term, *K*_*P*_*e*(*t*), is the proportional term, the second term is the integral control, and the third term is the derivative element; the constants *K*_*P*_, *K*_*I*_ and *K*_*D*_, all positive, are their respective gains. An improvement of the PID controller may be achieved by directly controlling 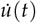 instead of *u*(*t*) [48]. Taking derivatives with respect to time in Eq. S35, the resulting control has the form

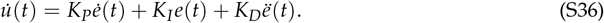

While the various terms in Eq. S36 are functions of continuous time *t*, they may have to be computed via discrete-time approximations. We use the following approximation due to its computational efficiency:

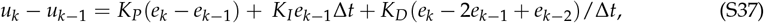

*k* = 1, 2, …, *K*_*f*_ ; here *e*_*k*_ := *e*(*k*Δ*t*), Δ*t* is a fixed sampling period, and *u*_*k*_ acts to approximate *u*(*k*Δ*t*). The range of *k* is *{*0, …, *K*_*f*_ *}* where *K*_*f*_ := [*t* _*f*_ /Δ*t*] is the largest integer not exceeding *t* _*f*_ /Δ*t, u*_0_ is a given initial condition, and it is assumed that *e*_*k*_, *k* = 0, 1, …, [*t* _*f*_ /Δ*t*] can be read from the system at time *k*Δ*t*. We applied the discrete-time PID controller defined in Eq. (S37) with Δ*t* = 0.01.

In applying the PID controller (Eq. S36) via the approximation defined by Eq. S37 to the epidemic social planner problem, we took the input control to be the employment relative to its prepandemic steady state, namely *u*(*t*) := *n*_*t*_ (see Eq. S10 with *n*_*SS*_ = 1), and the system’s output *y*(*t*) to be the effective reproduction number of the epidemic, ℛ _*e*_(*t*). The choice of *u*(*t*) is reasonable since it provides the planner with a direct policy control influencing both the mortality rate and welfare loss. Regarding the choice of the system’s output, simulation-based evidence has shown that according to the open-loop optimal control, ℛ _*e*_(*t*) converges to and then maintains a near-constant value slightly less than 1, for a significant part of the simulation horizon.

The configuration of the feedback system is depicted in Figure S3. Note that the PID controller is not used directly to shape the input *u*(*t*) of the econo-epidemic system, but rather serves to compute *u*_*c*_(*t*) from the tracking-error signal of the econo-epidemic system, *e*(*t*) := *r*(*t*) − *y*(*t*). This computation, performed in real time, is defined by Eq. S36 and approximated by Eq. S37. The resulting signal *u*_*c*_(*t*) typically is continuous, or at least piecewise continuous, in contrast with the input to the econo-epidemic system, *u*(*t*), which has to satisfy the aforementioned piecewise-constant constraint. This constraint is ensured by the controller in the lower loop of the figure, marked as the “secondary controller”. It is a time-varying system with two inputs, the tracking-error signal *e*(*t*) := *r*(*t*) − ℛ_*e*_(*t*) as well as the input-error signal *u*(*t*) − *u*_*c*_(*t*), and a single output, *u*(*t*_+_).

##### Definition of the secondary controller

The purpose of the secondary controller is to ensure that the input signal to the econo-epidemic system, *u*(*t*), satisfies the piecewise-constant constraint. To describe its dynamics, we denote by *P*_*j*_, *j* = 1, 2, …, the *j*^*th*^ maximal time-interval (period) during which *u*(·) has a constant value, labeled a *constant-value period*. Let *t*_*j*_ ∈ [0, *t* _*f*_) denote the starting time of *P*_*j*_, and observe that *P*_*j*_ = [*t*_*j*_, *t*_*j*+1_]. The constant value of *u*(*t*) throughout *t* ∈ *P*_*j*_, and the end time-point of *P*_*j*_, *t*_*j*+1_, are defined as follows. Given a *t*-dependent function *E*_*u*_(*t*) > 0, a constant *E*_*τ*_ > 0, and a *t*-dependent function *E*_*d*_(*t*) > 0, respectively labeled as *input-error threshold, tracking-error threshold*, and *dwell-time threshold*. Consider the starting time of *P*_*j*_, *t*_*j*_, for some *j* = 1, 2, …, such that *t*_*j*_ < *t* _*f*_. Recall that the PID controller runs in real time (or, better to say, approximates a real-time computation by using a given finite grid). We set *u*(*t*_*j*_) = *u*_*c*_(*t*_*j*_), which determines that *u*(*t*) ≡ *u*_*c*_(*t*_*j*_) ∀*t* ∈ *P*_*j*_. Meanwhile, *u*_*c*_(*t*) keeps on changing according to the computations by the PID controller. The constant-value period, *P*_*j*_, is terminated in response to one of the following two events, whichever occurs first:

For *t* ∈ *P*_*j*_,

1. |*u*(*t*) − *u*_*c*_(*t*)| ≥ *E*_*u*_(*t*) and |*e*(*t*)| ≥ *E*_*τ*_ and *t* − *t*_*j*_ ≥ *E*_*d*_(*t*),
2. *t* = *t* _*f*_.

In case of Event 1, we set *t*_*j*+1_ = *t* and reset *u*(*t*_*j*+1_) = *u*_*c*_(*t*_*j*+1_), while in case of Event 2, we set *t*_*j*+1_ = *t* _*f*_. Observe that the input error *u*(*t*) − *u*_*c*_(*t*) is reset to zero at the starting time of every constant-value period *P*_*j*_, *j* = 1, 2, ….

We chose the time-dependent function *E*_*u*_(*t*) to be reset at the start of every constant value period *P*_*j*_, namely at time *t*_*j*_, to a given base value *b*_*u*_ := *E*_*u*_(0) that is independent of *j* = 1, 2, Thereafter *E*_*u*_(·) is monotone decreasing during an early part of *P*_*j*_, and monotone non-increasing throughout *P*_*j*_, thereby guaranteeing that *E*_*u*_(*t*) jumps upwards to the value *b*_*u*_ at every time *t*_*j*_, *j* = 1, 2, This form of the input-error threshold *E*_*u*_(·) is designed to require larger input errors for terminating *P*_*j*_ sooner rather than later after its starting time. This, in turn, would tend to limit from below the dwell times (lengths) of constant-value periods hence potentially limiting from above their total number throughout the interval *t* ∈ [0, *t* _*f*_]. On the other hand, the time-dependent function *E*_*d*_(*t*) was chosen to be monotone increasing throughout the interval [0, *t* _*f*_], and have no resets. This was designed to permit more frequent switchings of constant periods early in the interval [0, *t* _*f*_] rather than late, which may be useful shortly after the outbreak of the pandemic, when the effects of modeling errors on loop signals can be large and costly as compared to later stages of the epidemic.

The specific functions *E*_*u*_(·) and *E*_*d*_(·) as well as the value of *E*_*τ*_ were chosen based on simulations of the system with various parameters. The resulting parameters of the PID controller are

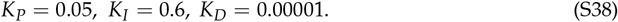

For the secondary-control parameters, the threshold function *E*_*u*_(·) has the form

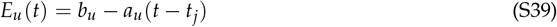

throughout the first part of *P*_*j*_, where *b*_*u*_ := *E*_*u*_(*t*_*j*_) is the base level at the start of *P*_*j*_ and *a*_*u*_ > 0 is a given constant. *E*_*u*_(·) is switched to a constant mode in the event that it reaches a given lower threshold level *θ*_*u*_ > 0, and it maintains that level until the end of *P*_*j*_. Thus,

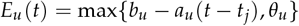

throughout *P*_*j*_, for given *b*_*u*_ > 0, *a*_*u*_ > 0, and *θ*_*u*_ > 0. We used the parameters *b*_*u*_ = 0.1, *a*_*u*_ = 0.001, and *θ*_*u*_ = 0.03, and *E*_*τ*_ = 0.03. Further, the threshold function *E*_*d*_(*t*), *t* ∈ [0, *t* _*f*_] is a affine function of the form

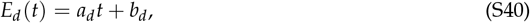

for a constant *a*_*d*_ > 0 and a base level *b*_*d*_ > 0. We chose *a*_*d*_ = 0.1 and *b*_*d*_ = 14.

The basic time unit for the problem under consideration is a day, and the end time is *t* _*f*_ = 630 days. We used the discretized PID controller defined by Eq. S37 with Δ*t* = 0, 01, hence it, performs a computation every 14.4 minutes. The secondary controller performs on the same schedule as the PID controller.

Regarding other quantities of the model, by Eqs. (S8) and (S9) the output of the econo-epidemic system is

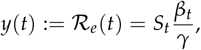

where *S*_*t*_ is the susceptible fraction of the population, *β*_*t*_ is the pandemic’s transmission rate defined by Eq. S10, and *γ*, the inverse of infectiousness period, taken at *γ* = 0.25 (see Supplementary Table S5). The total duration of the simulated epidemic is *t* _*f*_ = 630 days. The simulation results, depicted in Figure 4, indicate a substantially better tracking of the feedback control over an open-loop control.

### C Calibration

To provide a recent, familiar baseline, we calibrate the model to fit the U.S. economy as impacted by COVID19 [49]. Throughout, we work in daily terms. We calibrate the baseline used for simulation, while noting that calibrated parameters represent a plausible benchmark for our exploration of social planning policies for future pandemics. As a result, our evaluation of optimal policies includes simulations in parameter regimes that depart from this benchmark – sometimes in substantial ways. We then evaluate the extent to which social planning policies function effectively in a broad range of parameter choices while evaluating the possibility that the planner is uncertain and likely incorrect in their estimation of parameters before designing policy rules.

#### C.1 Calibration of the Epidemiological Model

Estimates of the latency period (1/*σ*) and the infectiousness period (1/*γ*) rely on studies from early in the COVID-19 pandemic [50, 51]. Their findings are confirmed by studies on infector-infectee pairs [52]. The Infection Fatality Rate (IFR), denoted *δ*, is based on estimates from the Imperial College COVID-19 Response Team [13] and a meta-analysis findings [6]. These sources estimate the IFR at 0.8%. The meta-analysis reports that the IFR of the disease across populations is 0.68% (0.53% − 0.82%), though it is noted that due to high heterogeneity, this might be an underestimate of the true IFR [6]. The typical duration of transitions from *I* to *D* is set at 11 days such that the average time between infection to death is 18 days, including both the incubation and infectious periods; we note that this period is consistent with but somewhat shorter than estimates for COVID-19 of closer to 21 days when including the distribution from infection to onset of symptoms and from symptoms to death [53].

#### C.2 Estimation-based Calibration of Transmission

We employ daily U.S. data to estimate key relations and use the point estimates to calibrate the model. The data series used are daily deaths, daily employment, lockdown measures, and the derived transmission rate. We estimate the equation for *β*_*t*_ :

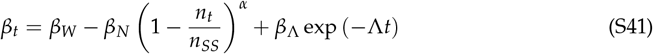

The results are shown in Table S2. The estimates imply the following. When 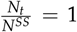 i.e., the economy is not locked and there are no sick or dead, at time *t* = 0:

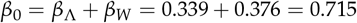

and so 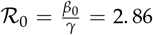 Given the estimated rate of decline, Λ = 0.12, in a little less than a month individuals adjust their behavior to the presence of the disease; subsequently, when exp (−Λ*t*) ≪ *β*_*W*_ we get:

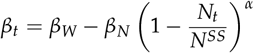

This implies *β*_*t*_ drops endogenously below *β*_*W*_ as a function of employment, yielding reproduction parameter ℛ_*t*_ variation between 1.5 and 0.8.

#### C.3 Calibration of the Economic Model

##### C.3.1 Discounting

We posit a 4% annual discount rate (*r* = 0.04), converted to daily terms (used by individuals and consequently by the social planner).

##### C.3.2 The value of *ϕ*

As assumed in prior work [54], we assume that anyone who has any symptoms self-isolates and does not work (*ϕ* = 1).

##### C.3.3 The endogenous response, lockdown policy, and employment

First, we follow the functional form proposed by previous work [11, 55] and postulate that the endogenous response function 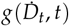 is given by:

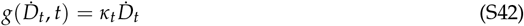

where

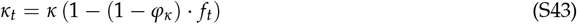

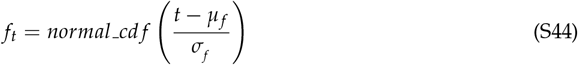

where *normal cd f* is the CDF of the normal distribution, *κ*_*t*_ is the time-varying parameter of the effects of the individual response on employment, and the parameters *µ*_*f*_, *σ*_*f*_, *ϕ*_𝓍_, and *κ* are estimated. The parameter *κ*_*t*_ express the idea that the endogenous individual response exhibits time decay *f*_*t*_. Next, we note that there is an overlap of compliance with lockdown and the endogenous response, so we use the maximal response as follows:

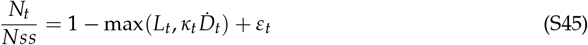

We non-linearly estimate equation S45 using U.S. data on 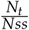,*L*, and 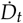 in the period from March 1, 2020 to February 28, 2021. The results are as follows:

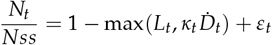

Finally, we take into account that without restrictions, employment would drop significantly. This can even reduce employment to levels below what is usually regarded as essential employment plus work from home. Empirical estimates for the U.S. indicate that the minimum employment level under the most stringent lockdown measures was around 0.65− 0.70 of full employment [56,57]. We therefore calibrate this level of employment to be 0.68 and set 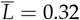.

##### C.3.4 Employment, Wages, and the Utility Function

For the planner problem and the simulations we further need to calibrate *w* and *A*. To do so we use two U.S. data points, as in a prior research [58]: the representative person earns an annual income of $58,000, using the 2019 estimate from the U.S. Bureau of Economic Analysis.

Thus, pre-epidemic, which we call steady-state (*SS*), when 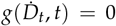, we get that the daily income *w* e is:

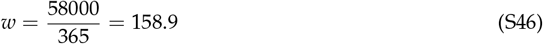

We set *A* = *w*.

##### C.3.5 Value of reduced mortality risk

The planner objective includes the term 𝒫^ss^ 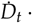 *VRMR*^*U*^, where *VRMR*^*U*^ is the value of reduced mortality risk. We determine its value and show how it fits in the social welfare function.

The central estimate for the monetary value of reduced mortality risk lost to COVID-19, *VRMR*^*USD*^, is 3.81 million USD, based on the EPA estimate of 270, 000 USD per year and an estimate of 14.1 years of remaining life on average [59].

To include these values in the social welfare function, we apply an oft-used methodology [60], as follows: denote the value of reduced mortality risk in utility terms by *VRMR*^*U*^, so that the event of death in the model is associated with utility loss of *VRMR*^*U*^. Individuals are indifferent between paying *SHARE*_*C*_ of their flow consumption and avoiding the risk *ε* of losing *VRMR*^*U*^, and not paying *SHARE*_*C*_ of their flow consumption and carrying the *ε* risk of losing *VRMR*^*U*^. Given the no-epidemic steady-state utility, this logic means that *VRMR*^*U*^ should satisfy the following indifference condition:

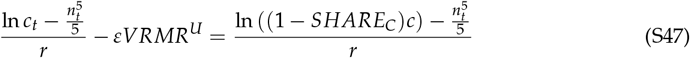

where

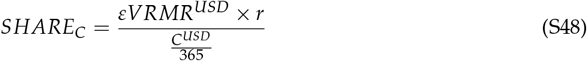

The representative agent would be willing to pay *SHARE*_*C*_ so as not to lose *VRMR*^*U*^ with an *ε* risk of death; the payment, *SHARE*_*C*_, is given by equation S48, paying *εVRMR*^*USD*^ × *r* each day, where *r* is the daily discounting rate is *r*.

Assuming *SHARE*_*C*_ ≪ 1 we get − ln (1− *SHARE*_*C*_) ≃ *SHARE*_*C*_ and using our modelling of *C* = *Y* (which we have taken to be 58, 000 USD), we get:

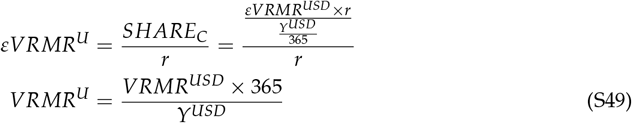

Thus the *VRMR*^*U*^ value we get is 23, 977 ≃ 24, 000 (also denoted *χ*) for the baseline *VRMR*^*USD*^ value of 3.81 million USD.

##### C.3.6 Vaccine Arrival Rate

The term *f* (*T*_*V*_) is the probability density function of the availability of a vaccine at time *T*_*V*_. This is an important term as it sets the horizon for the problem, acting as a hazard rate for leaving the state of the pandemic. It is an expression of the essential risk and uncertainty embodied in the planner problem. Note that were we to model an arrival time known with certainty, not only would an important real world aspect be removed, but such modelling might create an artifact in the optimal plan. The planner may enable an outbreak shortly before vaccine arrival, relying on the vaccine to eradicate it. Such a plan is not robust to delays in the arrival time. Relative to the interest rate *r*, expressing time preference, *f* (*T*_*V*_) plays the major quantitative role in discounting future streams.

We assume the Gumbel distribution, justified by the following logic. We assume that the arrival of the vaccine is a result of simultaneous competition among many firms. The time of arrival is the minimum development time across these firms. Note that over the course of 2020-2022 over 110 vaccines were in clinical trials and dozens more in pre-clinical evaluations. The distribution of arrival time is then well approximated by a Gumbel distribution [61], which is a member of the family of extreme value distributions. Specifically, it is used for modeling the minimum of a sample from many distributions, including exponential, logistic, and normal distributions. Under mild regularity conditions, it is suitable to be a model for a sample minimum even when the distributions from which the sample is drawn are unknown. In our setting, we remain agnostic about the distributions of vaccine development time by individual firms.

In terms of the model, *T*_*V*_ refers to the time of sufficient vaccination. With logistics, production times, gradual take-up rates, etc. an ex-ante expected 540 days seems reasonable relative to the March 2020 start date of the epidemic in the U.S.

The cumulative distribution function *G* (*x*) of a Gumbel distribution is defined over the real numbers and parametrized by a location parameter *µ*_*G*_ and a scale parameter *σ*_*G*_ :

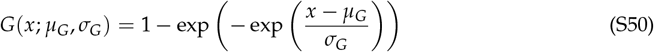

We anchor the distribution’s parameters (*µ*_*G*_, *σ*_*G*_), by positing that the mean of the distribution is 540 days, and that the probability of sufficient vaccination before day 360 is only 1%. These assumptions engender two linear equations:

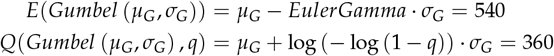

where *E* is the mean and *Q* is the quantile function. Targeting a mean of 540 and *Q*(*q* = 0.01) = 360 leads to the solution of *µ*_*G*_ = 565.83, *σ*_*G*_ = 44.74.

### D Supplementary Tables

**Table S1:** Econo-epidemic functions to coupled vaccine arrival, employment, endogenous response, and constraints.

| function | description | formal representation |
| --- | --- | --- |
| $f(T_V)$ | Gumbel distribution for vaccine arrival | $f(T_V; \mu_{T_V}, \sigma_{T_V}) = 1 - \left[ \exp \left( - \exp \left( \frac{x - \mu_{T_V}}{\sigma_{T_V}} \right) \right) \right]$ |
| $b(\dot{D}_t, \bar{L}, t)$ | employment response | $\max(\kappa_t \dot{D}_t, \bar{L})$ |
| $\kappa_t \dot{D}_t$ | endogenous response | $\kappa_t \dot{D}_t = \kappa (1 - (1 - \varphi_\kappa) \cdot f_t) \dot{D}_t$ |
| | | $f_t = \text{normal\_cdf} \left( \frac{t - \mu_f}{\sigma_f} \right)$ |
| $L_t$ | lockdown policy constraints | $\bar{L} \geq L_t \geq 0$ |

**Table S2:** Estimates of transmission relevant parameters given model fits, such that *R*_2_ = 0.74, *RMSE* = 0.0307, and *n* = 351 – complete details of model fitting are available in [49]. Elements are point estimates with standard errors in parentheses, and significance noted as * *p* < 0.10,** *p* < 0.05,*** *p* < 0.01.

| $\beta_\Lambda$ | $\Lambda$ | $\beta_W$ | $\beta_N$ | $\alpha$ |
| --- | --- | --- | --- | --- |
| 0.339***<br>(0.05) | 0.12***<br>(0.01) | 0.376***<br>(0.05) | 0.53***<br>(0.12) | 0.69<br>(0.31) |

**Table S3:** Parameters for the PID control algorithm. Data presented here was mentioned following the discussion of the PID algorithm (the space between equations S38 and S40, inclusive) and it is provided here in order to complete the summary discussion.

| Parameter | Description | Numerical value |
| --- | --- | --- |
| $E_u(t)$ | Input error threshold | $\max\{0.1 - 0.001(t - t_j), 0.03\}$, $t \in P_j$,<br>for every constant period $P_j$, $j = 1, 2, \dots$ |
| $E_d(t)$ | Dwell time time threshold | $0.1t + 14$ |
| $E_\tau$ | Tracking error threshold | 0.03 |
| $K_P$ | Proportional control gain | 0.05 |
| $K_I$ | Integral control gain | 0.6 |
| $K_D$ | Derivative control gain | 0.00001 |

**Table S4:**
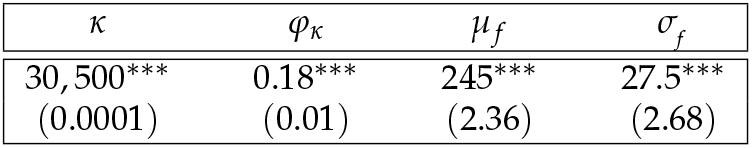
Economic model parameterization based on U.S. data during the period of March 1, 2020-February 28, 2021. The tables report point estimates with standard errors in parameters in parentheses, noting that *R*_2_ = 0.99, *RMSE* = 0.0139, and *n* = 365. Significance levels are noted as * *p* < 0.10,** *p* < 0.05,*** *p* < 0.01. The complete model fits are available in [49].

**Table S5:** Estimated disease parameters used in the econo-epidemic model derived in part from [49]. Note that the text uses the variables *t* _*f*_ and *T* interchangeably to denote the planner’s horizon.

| parameter | description | numerical value | source |
| --- | --- | --- | --- |
| $r$ | daily interest rate | $\frac{0.04}{365}$ | prevalent assumption |
| $\phi$ | fraction of ill not working | 1 | assumption |
| $\chi$ | weight assigned to death flows | 24,000 | VRMR est. |
| $\gamma$ | inverse of infectiousness period | $\frac{1}{4}$ | epi lit |
| $\sigma$ | inverse of latent period | $\frac{1}{3}$ | epi lit |
| $\delta$ | death rate | 0.008 | clinical lit |
| $\theta$ | inverse of period $I$ to $D$ | $\frac{1}{11}$ | clinical lit |
| $\beta_W$ | baseline transmission rate | 0.376 | est. |
| $\beta_n$ | employment effects scale parameter | 0.53 | est. |
| $\alpha$ | power of employment effects function | 0.69 | est. |
| $\beta_\Lambda$ | time effects scale parameter | 0.339 | est. |
| $\Lambda$ | exponential parameter in time effects | 0.12 | est. |
| $w = A$ | daily wages, productivity | 158.9 | data |
| $\kappa$ | scale parameter endogenous response | 30,500 | est. |
| $\varphi_\kappa$ | parameter endogenous response | 0.18 | est. |
| $\mu_f$ | mean of fatigue function | 245 | est. |
| $\sigma_f$ | std of fatigue function | 27.5 | est. |
| $\mu_{T_V}$ | location parameter of Gumbel distribution | 565.83 | est. |
| $\sigma_{T_V}$ | scale parameter of Gumbel distribution | 44.74 | est. |
| $\bar{L}$ | maximum restriction of economy | 0.32 | epi lit |
| $T$ | planner's horizon | 630 | |

### E. Supplementary Figures

**Figure S1:**
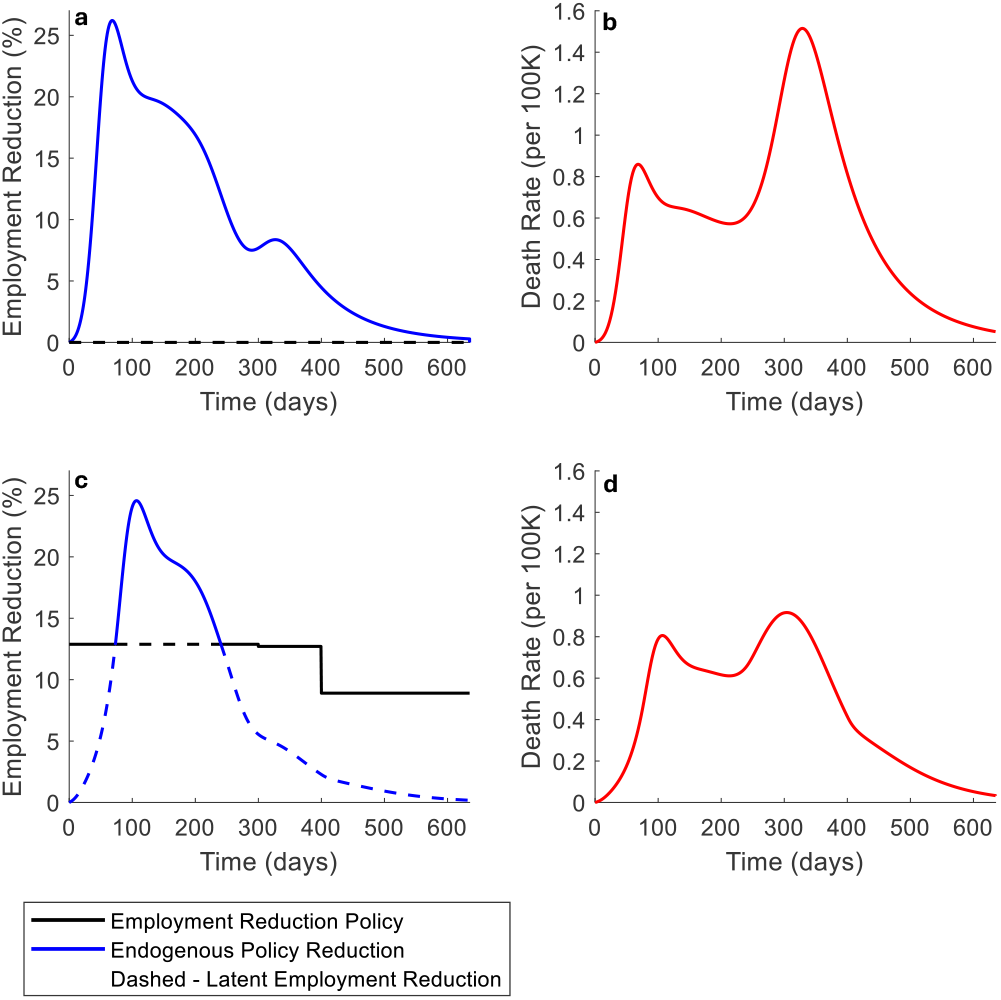
Baseline outbreak dynamics, endogenous response, and fatalities in the absence of optimal, policy-driven employment reduction. Dynamics depict cases of ℛ_<sub>0</sub>_ = 3.156 in a no employment reduction context (panels (a) and (b)) and with relaxed employment reduction (panels (c) and (d)). In both context, as death rates rise, individuals reduce interactions, sometimes more stringently than government-imposed employment reduction (whenever the blue curve is higher than the black curve). In (a)-(b), no reduction policy is imposed, leading to rapid spread and increasing deaths until the population reacts. In (c)-(d), the imposed restrictions are insufficient, prompting tighter individual measures as death rates escalate. Over time, the anticipated arrival of vaccines weakens the individual response, evident in both scenarios, where later larger death spikes elicit a diminished endogenous reaction. The cumulative fatalities per 100,000 in the two scenarios are 378.08 per 100,000 in case (a)-(b) and 280.02 per 100,000 in case (c)-(d).

**Figure S2:**
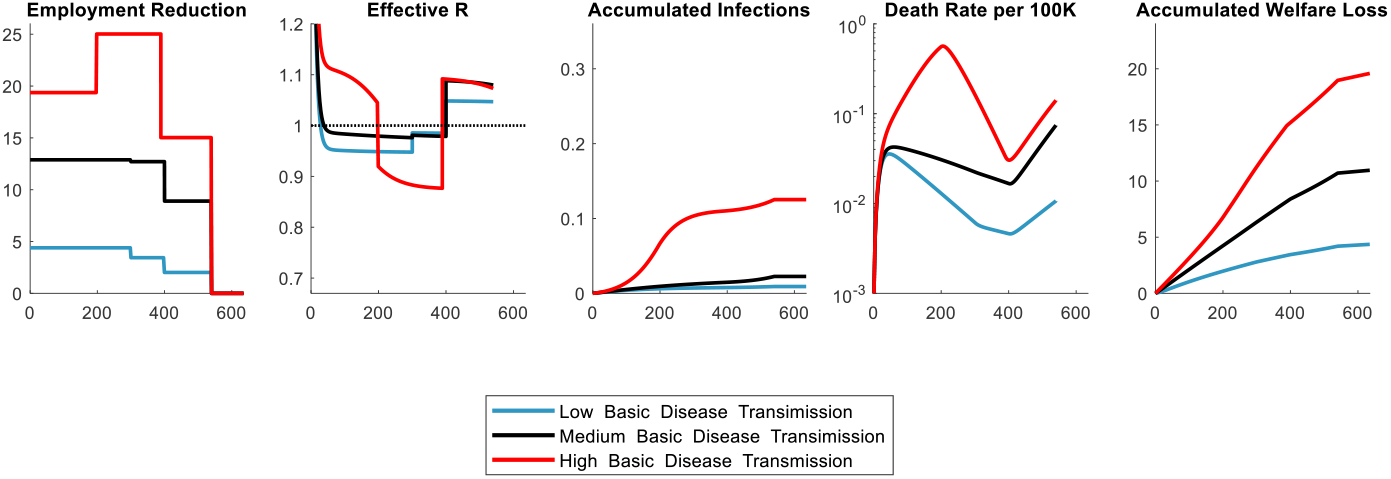
Econo-epidemic dynamics given optimal stepwise response policies. The policies outlined represent the optimal stepwise approach for basic transmission rates (*β*_w_) of 0.3,0.376, and 0.45, respectively for low, medium and high cases assuming accurate knowledge of disease parameters. As the basic transmission rate increases, the disease spreads more rapidly, necessitating a more stringent optimal policy. However, even with these measures, increased rates of infection and mortality may still occur as the basic disease transmission rises, due to the severe economic repercussions of overly aggressive employment reductions. The optimal policy appears to maintain the effective reproductive number (ℛ_eff_)close to 1.

**Figure S3:**
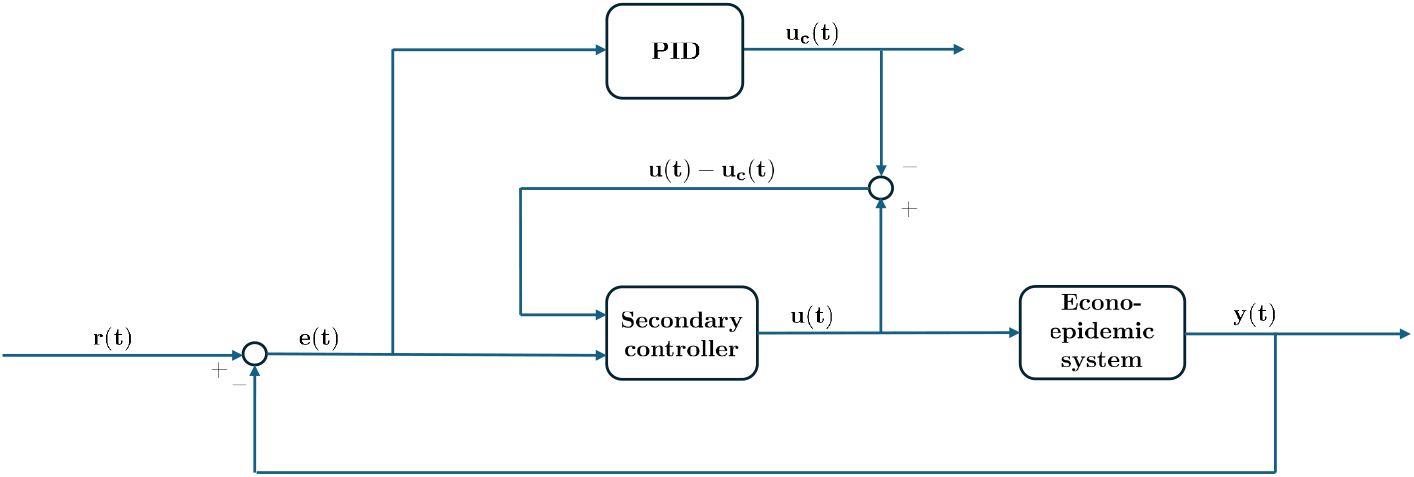
The relationship between the feedback controller and the controlled system.

**Figure S4:**
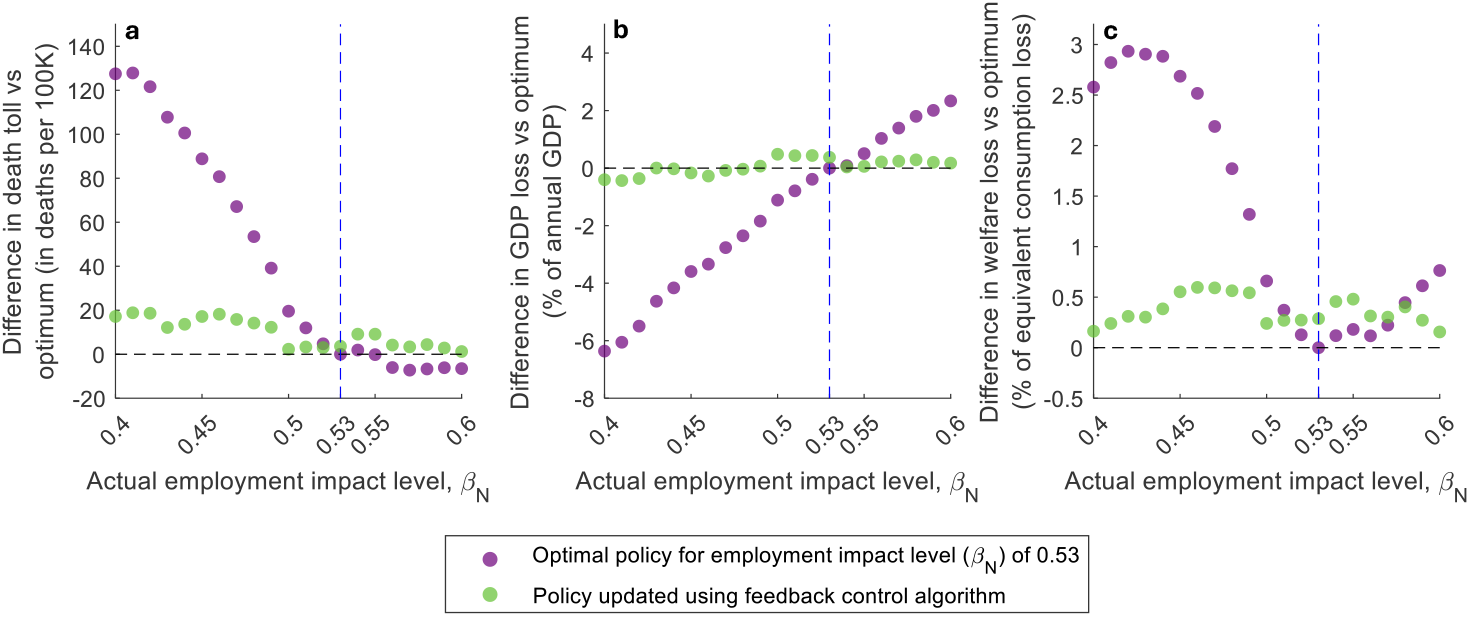
This Figure presents similar results to as in Figure 4, but with respect to variations in *β*_N_ instead of variations in *β*_W_ The optimal policy is sensitive to misspecification of the impact of employment on the spread of the disease, whereas the feedback policy approach is not The policy using feedback control maintains death toll (a), GDP loss (b) and WL (c) in levels that are close to optimal across all levels of employment impact.

